# Post-progression survival in advanced non-small cell lung cancer treated with anti-PD-1/PDL-1 monotherapy: progression after durable clinical benefit versus primary resistance

**DOI:** 10.1101/2023.03.09.23287023

**Authors:** Ivan Pourmir, Reza Elaidi, Zineb Maaradji, Hortense de Saint Basile, Monivann Ung, Benoit Gazeau, Laure Fournier, Bastien Rance, Laure Gibault, Mohammed Ismaili, Elizabeth Fabre

## Abstract

**Aims:** Anti-PD-1/PD-L1 can face the issue of tumor resistance in advanced non-small cell lung cancer (aNSCLC), mostly as primary resistance to the treatment. Still, disease progression (PD) also occurs after prior durable clinical benefit (DCB). Comparison of both situations and tools to evaluate residual benefit of PD-1/PD-L1 blockade are needed to understand and manage resistance.

**Methods:** We reviewed aNSCLCs with anti-PD-1/PD-L1 monotherapy, and disease progression per RECIST 1.1 (PD) in our center. Primary objective was comparison of post-progression survival (PPS) in primary resistance versus PD after DCB. Secondary objectives were to characterize patterns of PD after DCB; assess the feasibility and relevance of Tumor Growth Rate (TGR) in PD after DCB.

**Results:** 148 patients were included. Median PPS were 5.2months (95% CI: 2.6-6.5) and 21.3months (95% CI: 18.5-36.3) (p<0.0001) for primary resistance and PD after DCB respectively. Multivariable hazard ratio for death in PD after DCB versus primary resistance was 0.14 (95% CI: 0.07-0.30). 76.7% of PD after DCB occurred in ≤3 lesions; 72.1% occurred only in preexisting lesions. TGR suggested a persistent benefit of PD-1/PD-L1 blockade in at least 44.2% of cases.

**Conclusions:** PD after DCB was an independent factor of longer post-progression survival, with specific patterns of progression. TGR is a promising tool to assess residual benefit of immunotherapy.

**Highlights:**

- Anti-PD-(L)1 immunotherapy still faces the problem of resistance in lung cancer
- Resistance can be classified as primary or secondary (acquired)
- Outcomes and mechanisms of both situations need to be characterized
- This unique comparative study reveals diverging survival and progression patterns
- Prior durable clinical benefit associates with longer post-progression survival
- This data benefits our understanding and management of resistance to anti-PD-(L)1

## Introduction

### 1. Epidemiology of non-small cell lung cancer

Lung cancer is currently the leading cause of cancer death in the world, non-small cell lung cancer (NSCLC) being the most frequent subtype.^1^ Advanced stages of the disease (aNSCLC) are associated with poor prognosis in historical cohorts: median survival was 3.3 months if untreated and 8.6 months under conventional chemotherapies.^2^

### 2. Current immunotherapies. Indications of anti-PD-1/PD-L1 monotherapy

Advances in cancer immunotherapy have significantly improved this situation. NSCLC is among the first cancers that benefited from immune checkpoint inhibitors (ICI), especially monoclonal antibodies blocking the Programmed Death-1/Programmed Death Ligand-1 axis (anti-PD-1/PD-L1), with profound and durable responses in some cases.^3,4^ These drugs have been first approved as monotherapy for aNSCLC in 2015.^5–7^ They were designed to block signaling pathways dampening the anti-tumor immune response, although the precise pharmacodynamics are still not comprehensively elucidated.^8,9^

### 3. Definitions for resistance to anti-PD1

As other cancer types responding to PD-1/PD-L1 blockade, some aNSCLC will experience durable clinical benefit (DCB) under anti-PD-1/PD-L1 monotherapy, defined as a ≥6 month duration of controlled disease per RECIST 1.1 by the Society of Immunotherapy of Cancer (SITC).^10^ Unfortunately, still a majority of cases display tumor progression after initiation of these treatments, leading to new challenges in defining optimal therapeutic strategies and understanding the underlying biological processes. In order to standardize studies, the SITC taskforce recently advised a classification for this outcome:^10^

- Primary resistance, if tumor progression per RECIST 1.1 occurs <6 month after anti-PD-1/PD-L1 initiation.
- Secondary resistance, if tumor progression occurs under anti-PD-1/PD-L1 treatment and after ≥6 month of clinical benefit from the start of the therapy.
- Late progression, if tumor progression occurs after anti-PD-1/PD-L1 discontinuation and after ≥6 month of clinical benefit from the start of the therapy.

### 4. Need to characterize progression after DCB (PD after DCB)

Secondary resistance and late progression are less often encountered than primary resistance, given that still a minority of cases exhibits prior durable clinical benefit and that ICI are still of relatively recent use in this setting.^4,11,12^ Hence, clinical and biological data about aNSCLC progression after durable clinical benefit is scarce and uses various definitions of resistance to immunotherapy.^10^ There is therefore a crucial need for characterizing PD after DCB, in order to adapt therapeutic strategy to the specificities of those patients.^13^

Treatment continuation beyond progression is increasingly attempted on the basis of the theoretical pharmacodynamics of immunotherapy, even in clinical trials.^14^ Tian et al. recently reported an association between PFS under anti-PD-1/PD-L1 regimen and post-progression survival (PPS) of aNSCLC, but this retrospective data was focused on patients treated beyond progression and only 4% of the cohort was under monotherapy.^15^ Yet, there is a lack of harmonization of practice based on objective data in this new clinical setting. In particular, there is a need for objective and feasible tools to assess residual benefit of anti-PD-1/PD-L1 beyond progression. Tumor Growth Rate (TGR) appears as a promising tool to measure the effect of treatment in NSCLC under anti-PD-1/PD-L1.^16,17^

Beside, clinical and radiological data about aNSCLC progression after durable clinical benefit under anti-PD-1/PD-L1 monotherapy is precious to decipher its biological mechanisms because it allows the observation and understanding of the resistance phenomenon in human individuals subject to pure PD-1/PD-L1 blockade. Indeed, latest approved regimens are combinations such as chemo-immunotherapy or anti-PD-1/anti-CTLA-4. Thus, looking at those patients, responses or progressions could be attributed to the combined drugs, precluding robust conclusions on anti-PD-1/PD-L1 escape.

In order to acquire better understanding of the specificities of PD after DCB and compare post-progression survival (PPS), we collected clinical and radiological data of aNSCLCs on anti-PD-1/PD-L1 monotherapy in our center. This enabled us, on the basis of SITC guidelines, to identify and characterize cases of primary resistance and PD after DCB. We then performed a comparative analysis of PPS between these groups and an in-depth characterization of PD after DCB.

## Materials and Methods

### 1. Cohort selection, exclusion criteria, radiological definitions

This monocentric, retrospective study included patients from European Georges Pompidou Hospital who started anti-PD-1/PD-L1 monotherapy between 30^th^ June 2015 and 31^th^ October 2019, and presented documented disease progression per RECIST 1.1 (PD) after at least 1 cycle of anti-PD-1/PD-L1.

Patients were excluded in case of other active neoplasm in the last 5 years; prior anti-PD-1/PD-L1 treatment course; concomitant immunosuppressive therapy (including systemic corticosteroid above 10mg per day); non-cancer related death before PD; lost to follow up before PD; insufficient baseline imaging data for subsequent disease assessment.

Included patients were classified into 2 groups:

- Anti-PD-1/PD-L1 primary resistance if PD occurred less than <6 months after anti-PD-1/PD-L1 initiation.
- Progression after DCB if PD occurred at least ≥6 months after anti-PD-1/PD-L1 initiation.

Within the PD after DCB group, patients were either categorized as oligoprogressive (OPD) if the number of progressive lesions per RECIST1.1 was ≤3 (including intracranial lesions), or categorized as non-OPD if >3 progressive lesions.^18,19^

### 2. Tumor kinetics evaluation by Tumor Growth Rate (TGR)

This exploratory analysis was run to evaluate the interest and feasibility of TGR, as an objective tool to assess remaining benefit of immunotherapy on tumor growth beyond PD after DCB. The global growth kinetics of cancer at a given date was estimated by the TGR, in percentage of global tumor volume increase per month. It was calculated from the sum of longest diameters of target lesions by RECIST 1.1, as previously published (Supplementary Fig. S1).^16,17^

TGR was assessed for patients of the PD after DCB group at baseline of ICI treatment and at PD, provided there was no change in antineoplastic treatments between the 2 consecutive imaging studies used to calculate the TGR. The TGR was also used to specifically estimate the growth kinetics of progressive lesions in OPD patients of the PD after DCB group. In this case it was calculated from measures of progressive lesions only.

### 3. Clinical, pathological and radiological data

Clinical, pathological and radiological data were extracted from electronic patient files.

PD-L1 expression in tumor tissue prior anti-PD-1/PD-L1 treatment was found in pathological reports and expressed as Tumor Proportion Score (TPS). TPS resulted from exploratory analyses run on some patients before 1^st^ line Pembrolizumab approval, and after that, from routine pathological evaluation as required from european and french guidelines.

Patients were followed with contrast-enhanced chest-abdominal-pelvic CT, and brain MRI when warranted. RECIST1.1 evaluation was collected from imaging reports for all patients. For the PD after DCB group, RECIST1.1 evaluation, OPD status assessment and measures were performed by a radiologist specialized in immunotherapy of lung cancer (Dr M. Ung) and a medical oncologist trained in radiological evaluation (Dr I. Pourmir). In this group, the consecutive imaging performed after the first PD assessment was also systematically reviewed.

Survival data was completed from the publically available national death register (Institut National de la Statistique et des Etudes Economiques). Study data was collected and managed using REDCap electronic data capture tools hosted at our institution.^20,21^

### 4. Statistical analyses

The primary objective was to compare PPS between aNSCLC patients with primary resistance versus PD after DCB on anti-PD-1/PD-L1 monotherapy. Secondary objectives were to analyze: PPS according to OPD status in the PD after DCB group; progression sites in primary resistance and PD after DCB groups. The endpoint PPS was defined as the time elapsed between progression time on ICI and death or last-contact.

Survival rates were estimated using the Kaplan-Meier method and statistical significance of post-survival differences used the Log-rank test. We used a Cox regression full model including the following covariables of interest to obtain hazard ratios (HR) with their 95% confidence intervals (95% CI): age at progression, sex, histology, treatment line of ICI, ECOG-PS (0-1 vs ≥ 2) at ICI initiation and at progression time, and disease stage (localized vs advanced/metastatic).

Characteristics of individuals were compared between the PD after DCB and primary resistance groups using a Wilcoxon rank sum test for numerical values and exact Fisher test for categorical values. PDL1 TPS categories were compared using a weighted global Fischer test.

Follow-up completion was assessed using Clarck’s index (Lancet, 2002).

Covariables effect sizes in multivariable model were statistically significant if their p-values were < 0.05. Statistical analyses were performed using R.4.1.3.

### 5. Ethics

The present study has been accepted and registered to the relevant institutional research and ethical committee (European Georges Pompidou Hospital); it has been conducted in accordance with the Declaration of Helsinki. All patients provided informed consent under the European Georges Pompidou Hospital approved protocol allowing collection and analysis of data.

## Results

### 1. Patient’s characteristics

Among the 189 patients who started a first course of anti-PD-1/PD-L1 monotherapy for aNSCLC from 30^th^ June 2015 through 31^th^ October 2019 were screened, 148 were included in the study, 41 were excluded for reasons detailed in study diagram (Fig. 1). Baseline characteristics of included patients are shown in Table 1. A total of 43 patients were classified into the PD after DCB groups and 105 patients in the primary resistance group, and then analyzed for the primary objective. As of PD, global median follow up index was respectively 0.99 (95% CI: 0.98-0.99) and 0.99 (95% CI: 0.44-0.99) in primary resistance and PD after DCB groups. PD-L1 score categories were balanced between primary resistance and PD after DCB groups although a substantial proportion of patients (42.6%) had no PD-L1 TPS evaluation prior to ICI available. Out of 15 patients with a known baseline TPS ≥50%, 9 (60%) received Pembrolizumab as their first line of treatment.

**Figure 1:**
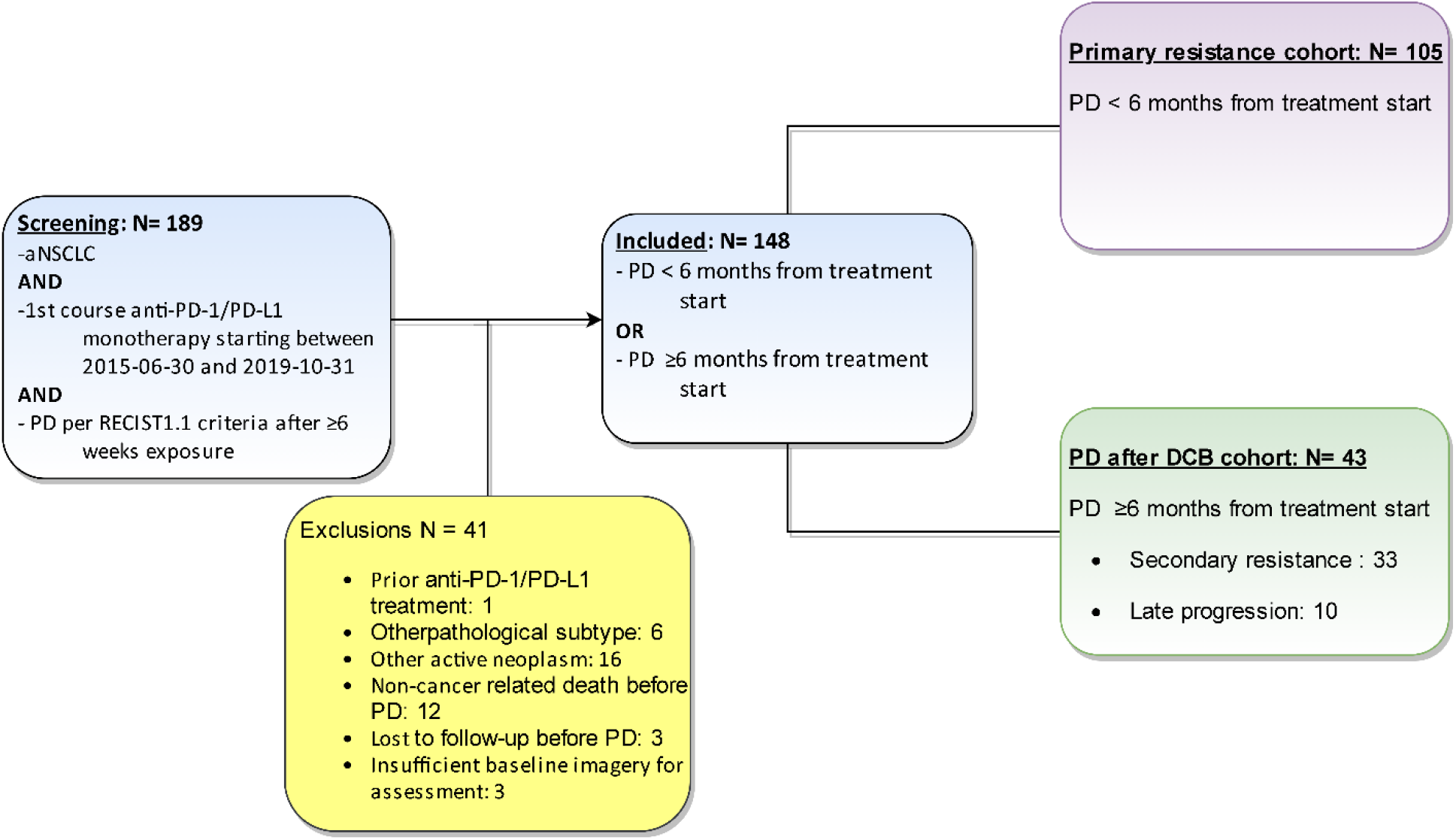
Study flowchart.

**Table 1:**
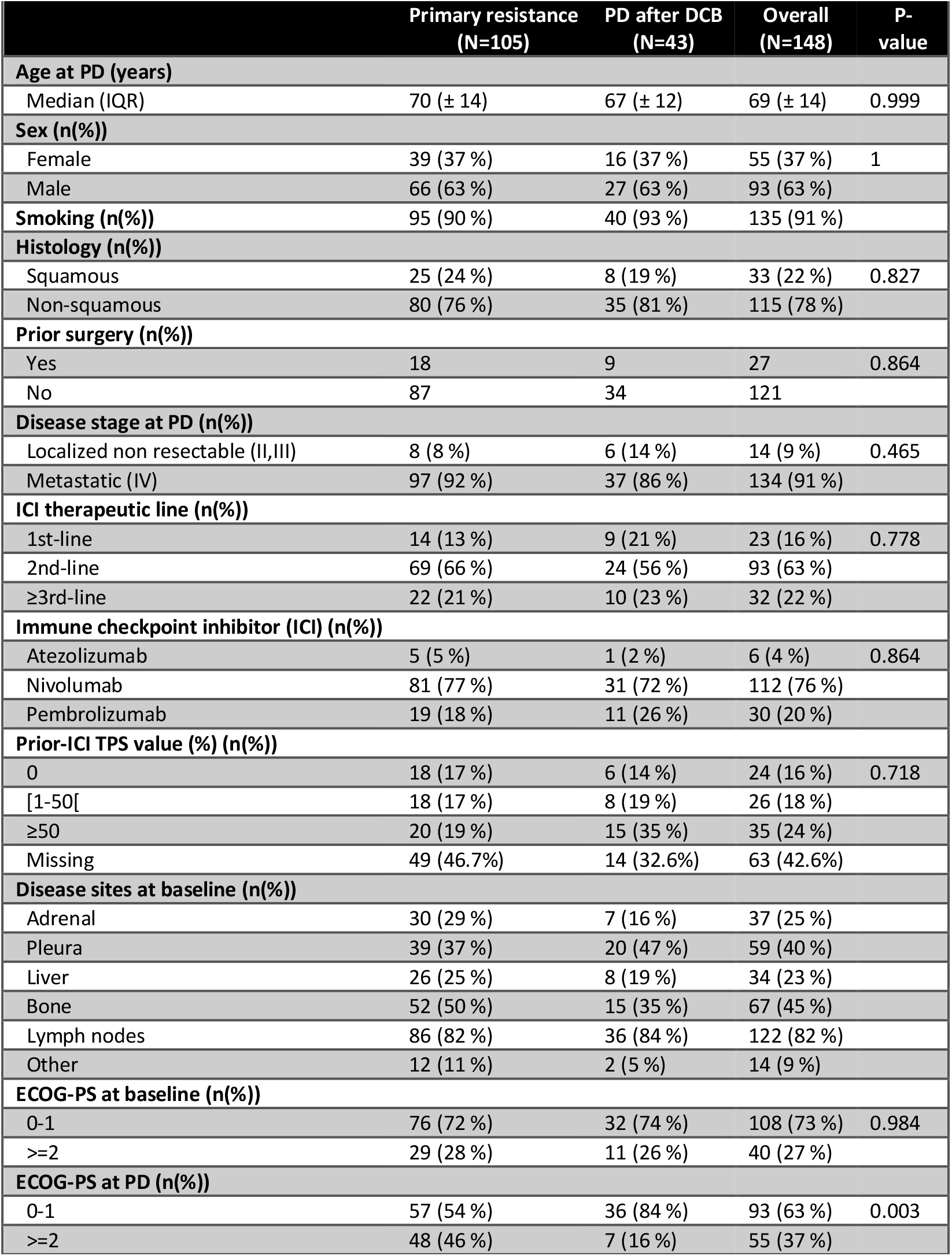
Patients characteristics. “at baseline”: at baseline of anti-PD-1/PD-L1 treatment. “at PD”: at first PD after anti-PD-1/PD-L1 initiation.

### 2. Comparative analysis of PPS in primary resistance and PD after DCB groups

Median PPS of aNSCLC treated with anti-PD-1/PD-L1 monotherapy was 5.2months for primary resistance (95% CI: 2.6-6.5) and 21.3months (95% CI: 18.5-36.3) (p<0.0001) for PD after DCB respectively (Fig. 2). Multivariable hazard ratio for death in PD after DCB versus primary resistance was 0.14 (95% CI: 0.07-0.30) (Fig. 3).

**Figure 2:**
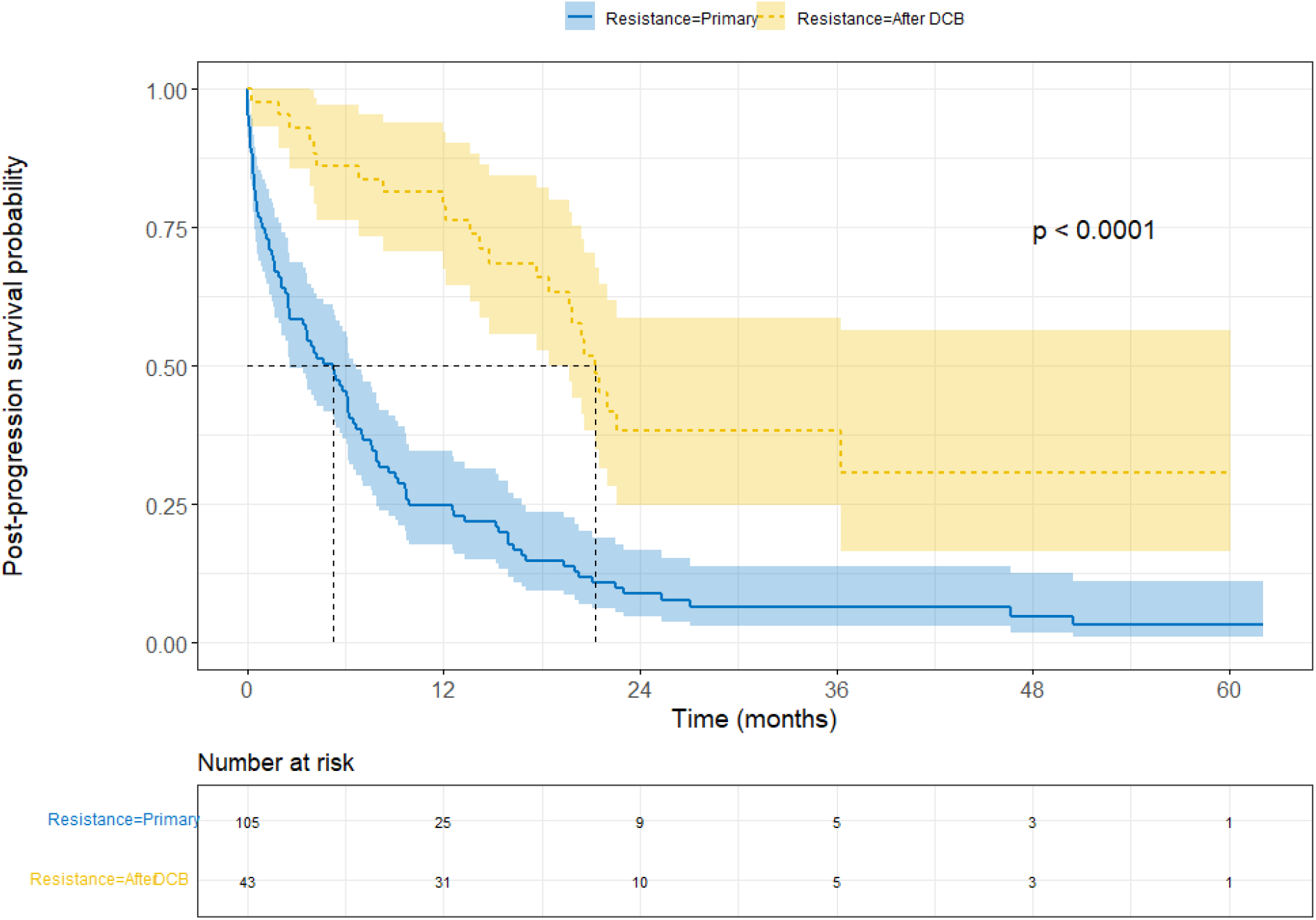
Comparison of PPS in PD after DCB versus primary Resistance. Kaplan– Meier survival curves showing PPS of primary resistance versus PD after DCB groups. P-value for Log-rank test.

**Figure 3:**
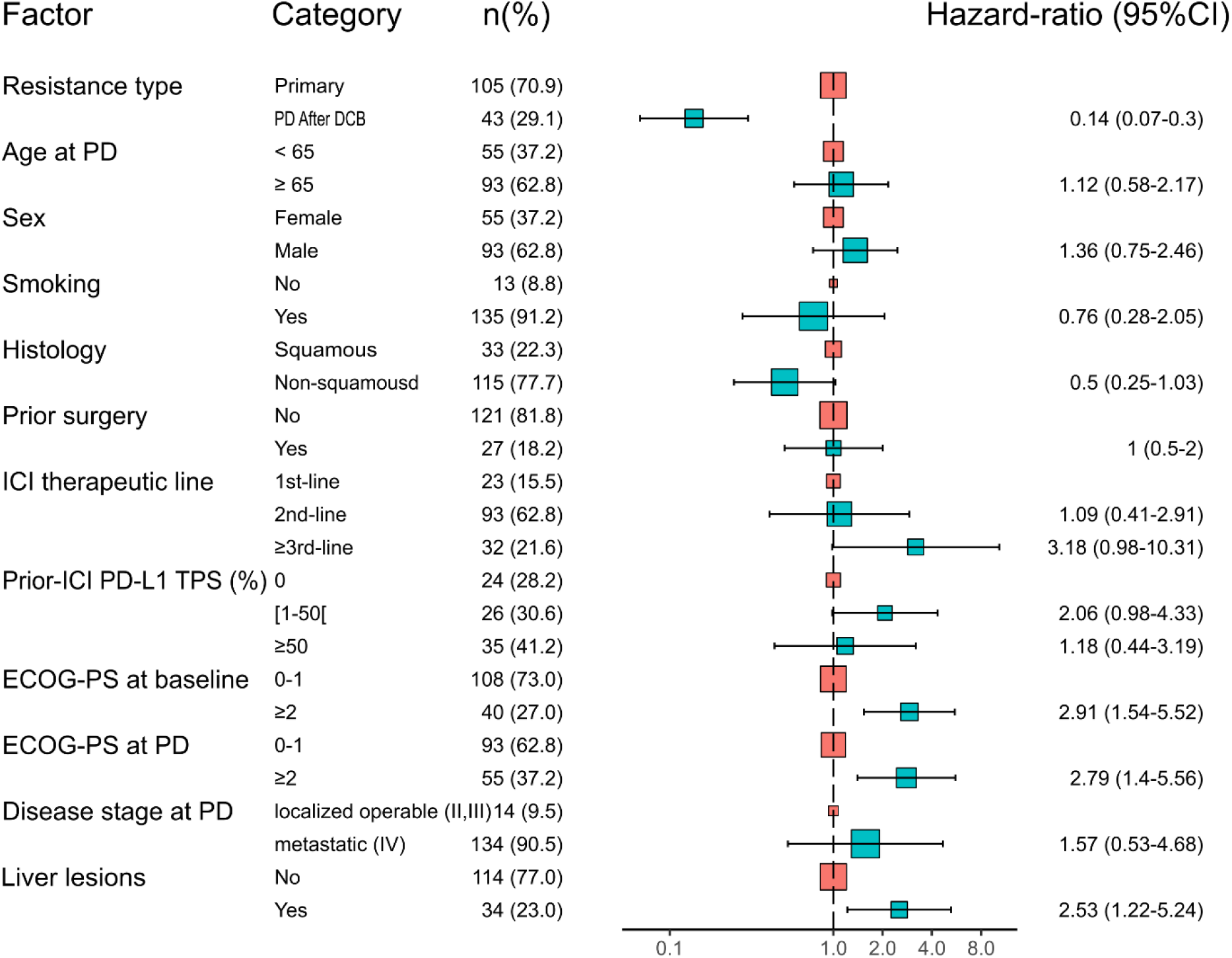
Cox multivariable model for PPS in PD after DCB versus primary Resistance. Forest plot showing hazard ratios for death in PD after DCB versus primary Resistance and other significant covariables.

### 3. Occurrence of PD after DCB

43 out of 61 (70.5%) aNSCLCs having DCB and meeting eligibility criteria eventually experienced PD, with a median time to progression of 11.4 months (95% CI: 9.4-16.4 months). Within the PD after DCB group, 33/43 (76.7%) patients were classified as secondary resistance in line with SITC criteria, 10/43 (23.3%) patients experienced PD after a break from therapy and met the criteria for being classified as late progression.

### 4. OPD prevalence in PD after DCB and association with PPS

Within the PD after DCB group, 33/43 (76.7%) patients had PD occurring in ≤3 lesions and were thereby classified as OPD, whereas 10/43 (23.3%) patients had PD occurring in >3 lesions and were classified as non-OPD. Most OPD patients (58%) displayed PD in only 1 lesion. Median PPS was 22.0 months (95% CI: 20.5-NR) and 19.2 months (95% CI, 12.2-NR) in OPD and non-OPD respectively, with univariable HR for death in OPD = 0.65 (95% CI: 0.27-1.57). Upon multivariable analysis including PDL-1 expression and PFS on anti-PD-1/PD-L1 (significant factors in full model), OPD was found to be a protective factor with a lower hazard for death after DCB compared to non-OPD: adjusted HR for death = 0.37 (95% CI: 0.09-1.59).

### 5. TGR evaluation in patients continuing PD-1/PD-L1 blockade beyond PD after DCB

An exploratory analysis of TGR was done in patients of the PD after DCB group having sufficient imaging data available. In at least 19/43 (44.2%) patients, TGR suggested ongoing benefit of anti-PD-1/PD-L1 treatment despite PD. For these patients, TGR assessed at PD was either null/negative, or inferior to TGR measured at baseline before ICI when both time points were available (Supplementary Table S1). Overall, the global kinetics of cancer growth explored by TGR in the PD after DCB group was low to moderate with a median volume growth of 2.1%/month (IQR 19.6%/month). In OPD assessable patients (n=24), median TGR measured specifically on progressive lesions was 24.6 %/month (IQR 17.8%/month) (Supplementary Table S2). Which is to say that, progressive lesions in OPD patients had a median volume growth of about one-quarter per month.

### 6. Sites of PD after DCB

Mains sites of PD for the PD after DCB group were lungs for 19/43 (44.2%), lymph nodes for 15/43 (34.9%) and central nervous system (CNS) for 5/43 (11.6%) patients. PD occurred exclusively in lymph nodes in 11/43 (25.6%) patients, consistent with previously published observations.^22^ At least 8 of these 11 cases showed no extra lymphatic progression at next imaging. In 31 out of 43 (72.1%) patients, progression occurred exclusively in preexisting lesions (i.e. lesions that had been already identified prior ICI treatment). There were no substantial differences regarding these patterns of progression between secondary resistance and late progression as defined by the SITC. Interestingly, spreading to previously non affected organs occurred more frequently in primary resistance than in the PD after DCB group (Table 2).

**Table 2:**
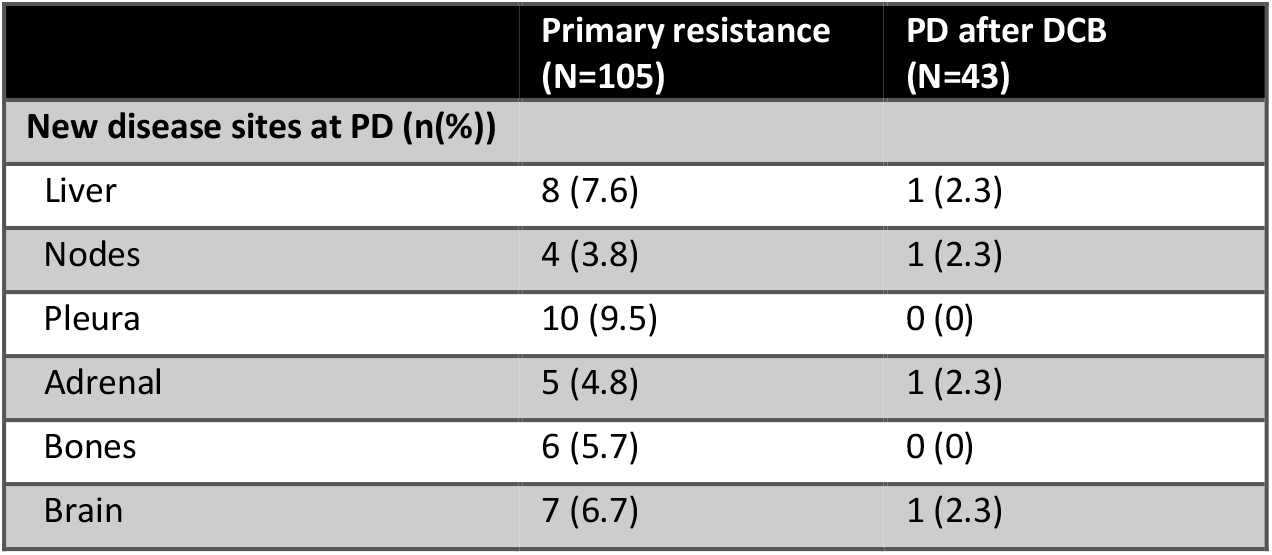
Tumor progression occurring in previously non affected organs.

|  | Primary resistance<br>(N=105) | PD after DCB<br>(N=43) |
| --- | --- | --- |
| **New disease sites at PD (n(%))** |  |  |
| Liver | 8 (7.6) | 1 (2.3) |
| Nodes | 4 (3.8) | 1 (2.3) |
| Pleura | 10 (9.5) | 0 (0) |
| Adrenal | 5 (4.8) | 1 (2.3) |
| Bones | 6 (5.7) | 0 (0) |
| Brain | 7 (6.7) | 1 (2.3) |

### 7. Disease spreading after OPD

24/28 (85.7%) radiologically assessable patients with OPD after DCB showed no new lesion appearance at next evaluation, with a median time of 2.38 months between PD imaging and the next imaging (Supplementary Table S2).

## Discussion

Progression appears as a frequent event even among patients experiencing DCB on anti-PD-1/PD-L1 monotherapy, it was found in 70.5% of those patients. This is to our knowledge the first study providing a comparative assessment of outcomes from aNSCLC progressing on anti-PD-1/PD-L1 monotherapy before or after DCB, as defined by the SITC. It confirms diverging outcomes and patterns of progression between both situations. This is also the first study to explore the feasibility and usefulness of TGR in the setting of progressive aNSCLC after DCB.

### 1. Management of PD after DCB

Our results suggest a drastic difference in PPS depending on whether or not PD occurs after DCB. They support preliminary data of non-comparative studies, suggesting that PD after DCB is a particular situation with dramatically different prognosis compared to primary resistance.^22–25^ This distinctive feature must be taken into account when discussing the management of PD after DCB for a given aNSCLC patient. In addition of extended PPS, most patients of the PD after DCB group (36/43 – 83.7%) had a Performance Status ≤1 at PD. These characteristics allow considering a broad scale of drug and non-drug treatments of NSCLC for subsequent treatment. However, among patients experiencing PD after DCB, the OPD and non-OPD categories must also be distinguished.^25^ Subsequent treatments received by patients of the primary resistance and PD after DCB groups are detailed in Supplementary Table S3.

OPD appears as the most frequent pattern of PD after DCB and as a good prognosis factor in this situation, consistent with other studies.^22–26^ OPD is theoretically defined as the development of a restricted number of tumor lesions.^18^ There is yet no consensus about a precise number of progressive lesions that should be used to categorize OPD or non-OPD patients. Notwithstanding, it is largely admitted that such a number should be based on possibilities of local treatment.^27^ For this reason the limit of ≤3 progressive lesions was chosen for our study, which is widely used in aNSCLC publications and reflects the practice in our center.^23,25,26^

The predominance of OPD status should lead to consider local treatments such as stereotactic body radiation therapy (SBRT), thermo-ablative therapy or even surgery.^28^ To date our study is the first to show that most OPD occurring after DCB experience no further spreading of aNSCL on the short/medium term. This important data strengthen the interest of local treatment. The estimation of TGR for OPD lesions provided in this study may also be useful to decide time and technic for such treatment. For instance, a given TGR of OPD lesions would prompt quick initiation of SBRT before the gross tumor volume exceeds the technical possibilities.

Furthermore, we have found, as other authors, that PD after DCB is frequently exclusively located in preexisting lesions.^22,25^ This should be taken into account to decide local treatment. Such a strategy has been shown as potentially beneficial in oncogene-addicted aNSCLC, but results of comparative prospective studies remain to be published.^29^ In spite of these theoretical advantages, a minority of PD after DCB (4/43 – 9.3%) was treated with local therapies in our cohort (Supplementary Table S3). This might be due, to some extent, to the lack of follow-up data available at that time for clinician to support such treatments in PD after DCB.

On the other hand, our study suggests a potential benefit of systemic immunotherapy continuation in case of PD after DCB through the assessment of TGR. This is for instance the case if a patient experiences PD after DCB, if the TGR, calculated with help of an additional imaging performed a couple of weeks later on ICI, is still inferior to the baseline TGR. It therefore suggests a persistent effect on tumor growth, in spite of PD. This makes ICI continuation and combination (with additional agents such as chemotherapies, targeted therapies, ICI, etc.) a promising option to be preferred to treatment line switch.^28^ This is to our knowledge the first published work to apply the TGR in the setting of progressive aNSCLC after DCB, although it has been used in other studies including NSCLC patients treated by ICI.^30,16,17,31^ TGR appears as a promising and feasible tool, given the current paucity of objective criteria to establish a persistent benefit of ICI after PD. It should be noted that TGR relies only on RECIST measures of target lesions. For this reason, a negative TGR can be observed although there is a tumor progression, for instance in non-target lesions. Nonetheless, this would still indicate persistent benefit, at least on the proportion of disease burden residing in target lesions.

### 2. Considerations for the mechanisms of PD after DCB and acquired resistance

Despite prior sustained controlled disease on anti-PD-1/PD-L1 monotherapy, aNSCLC still end up progressing in the vast majority of patients. Mechanisms of resistance to anti-PD1/L1 immunotherapy still need to be elucidated, although multiple hypothesis based on preclinical models have been formulated.^32^ Resistance after DCB or secondary resistance could rely on distinct but also common mechanisms to primary resistance.^33^ In this context, implementation of clinical, pathological and biological data gathered in cohorts of patients treated with anti-PD1/L1 monotherapy seems pivotal to select and delve into the most relevant of these hypotheses.

#### 2.1. Sites of progression

Preferential sites of progression after DCB could indicate specificities of the microenvironment favoring a delayed immune escape. In the PD after DCB group, most frequent progressive sites were lungs, lymph nodes and CNS. The same hierarchy was observed in secondary resistance and late progression subgroups as defined by SITC. A similar repartition have also been reported by Heo et al. in a cohort of aNSCLC mostly treated with anti-PD-1/PD-L1 monotherapy and using a comparable definition of secondary resistance.^22^ It has been hypothesized on the basis of preclinical models that lung microenvironment could be prone to secondary resistance.^34–36^ Clinical data presented here is consistent with this hypothesis. Interestingly, disease spread to new organs was more frequent in primary resistance than in PD after DCB (Table 2).

#### 2.2. Lymphatic progression after durable clinical benefit is a distinct entity

As previously reported, a substantial proportion (25.6%) of PD after DCB occurred only in lymph nodes.^22^ It could be objected that these lesions simply witness a subclinical growth of the disease in neighboring organs. Nevertheless, the observation in our study, that the majority of these PD showed no involvement of other organs at next imaging, supports the theory that a secondary resistance limited to lymph nodes is a distinct biological entity. It has been hypothesized that this could be due to long term effects of PD-1/PD-L1 blockade on intra-lymphatic antitumor immune cells, either on their survival or on chemokine signals enabling their location in nodes.^23,26^

#### 2.3. PD after DCB is a localized phenomenon arising in preexisting lesions

The majority of cases of PD after DCB in our study occurred as OPD in preexisting lesions, as reported in previous studies.^25,24,22,37^ Here again, the biological relevance of our observations is strengthened by the careful review of imaging performed after the first PD assessment. This suggests sustainability of the systemic immune surveillance under anti-PD-1/PD-L1 monotherapy, along with the localized evolution of few preexisting lesions toward immune evasion or immune suppression. This localized resistance could arise for instance from cancer cells immunoediting, or from the development of immunosuppressive conditions within a given lesion.^33^ Furthermore, PD after DCB is less prone to arise in new organs compared to primary resistance. This might indicate an evolution of aNSCLC under long-term PD-1/PD-L1 blockade toward a better fitness in already invaded tissue, rather than the colonization of completely new tumor microenvironments.

### 3. Other considerations about strengths and limitations of this study

In contrast with other publications using non-consensual and heterogeneous criteria, we used the definitions provided by the SITC, in order to contribute to the necessary harmonization of clinical and translational research about immunotherapy resistance. This would hopefully help collecting homogeneous data in the future and performing aggregative and reproducible analyses.

It is worth noting that our PD after DCB group comprised the 2 subgroups of secondary resistance (n=33) and late progression (n=10) that should occasionally be differentiated. Indeed, aNSCLC progression in the setting of late progression could rely on different mechanisms from secondary resistance, given that it would happen off therapy. For instance, late progression could be due to an impaired global immune surveillance after PD-1/PD-L1 blockade discontinuation and would indicate that the dysfunction of antitumor immune cells is not definitively reversed by anti-PD-1/PD-L1 monotherapy. However, although based on small numbers, there were no striking differences in the patterns of progression between these two subgroups.

Nevertheless, definition of secondary resistance by the SITC might also be criticized, particularly in view of the 6 months DCB thresholds chosen to differentiate it from primary resistance. As explained by SITC experts, this minimal period of DCB is necessary to exclude aNSCLC growing slowly under ICI treatment with no prior benefit, but classified as stable disease (SD) per RECIST1.^10^. On the other hand, such thresholds may misclassify some aNSCLC as primary resistant, although they have showed prior response to immunotherapy before PD and might be relevantly categorized as secondary resistant from a biological point of view. Schoenfeld and colleagues have recently challenged these definitions and demand their adaptation.^38^ This controversy also illustrates the interest of TGR as a tool to objectively identify patients that experience some benefit early on ICI treatment (i.e. those who have a reduction in TGR, even if evaluated as SD by RECIST1.) prior to resistance, and improve secondary resistance definition. This approach has been used by Ferté et al. to evaluate benefit of anticancer drugs in early phase clinical trials data^39^. However, due to the retrospective design of our study, TGR could not be analyzed in all patients, which prevented us to include this parameter in multivariable analyses.

Our study provides homogeneous data, as we only included patients under anti-PD-1/PD-L1 monotherapy. This fact increases, among other things, the relevance of biological hypotheses about secondary resistance based on this data. Conversely, some previous publications pooled anti-PD-1/PD-L1 monotherapy with combinations including anti-CTLA-4, chemotherapy or tyrosine kinase inhibitors. For instance, Schoenfeld et al. analyzed a cohort of patients treated either with anti-PD-1/PD-L1 monotherapy or combined with anti-CTLA-4, assuming that patterns and mechanisms of resistance should be similar between both types of therapies but without providing a subgroup analysis.^25^ In addition, our cohort consists mostly of Caucasian patients with data that might biologically better apply to population of western countries. For instance, one of the largest cohort of secondary resistance of aNSCLC (n=51 patients under anti-PD-1/PD-L1 monotherapy) originates from Korea and comprises a high proportion of EGFR-mutated NSCLC (≥17.4%).^22^

In addition, our imaging review of PD after DCB, performed by two trained investigators, improves the confidence in patterns of progression that we demonstrated here. Indeed, these patterns could have been otherwise confounded by the interobserver variability of imaging, or insufficient sensitivity of morphological imaging. However, the prevalence of cancer progression in the CNS can be underestimated by the limitations and non-systematic use of brain imaging for the follow-up of aNSCLC without previous history of CNS metastasis in our retrospective study.

Also, a substantial proportion of patients (42.6%) had no PD-L1 TPS evaluation prior to ICI available. This is explained by the fact that a large share of our cohort received Nivolumab as ≥2nd line treatment which was indicated regardless of TPS, before 1st line Pembrolizumab approval. Therefore, PD-L1 scoring was not routine practice at that time. To our knowledge, all other publications on this topic face the same issue, having many TPS value missing or not providing them at all.^22,24–26,37^ However, some exploratory pathological analyses of PD-L1 performed in our center enabled us providing TPS values even for some of these ≥2 line Nivolumab patients.

We provide here the first real life comparative study of PPS between primary resistance and resistance after DCB in aNSCLC treated with immunotherapy. PD after DCB under anti-PD-1/PD-L1 monotherapy is shown to be an independent factor of extended PPS and displays distinct patterns. As previously reported, OPD status tends to associate with better prognosis among PD after DCB patients. TGR is a promising tool to guide decision of ICI continuation and to refine secondary resistance definition. Altogether this data shall contribute to better basic understanding and clinical management of aNSCLC’s secondary resistance to immunotherapy (Fig. 4). It will profitably add up with other studies and improve significance with larger numbers of observations.

**Figure 4:**
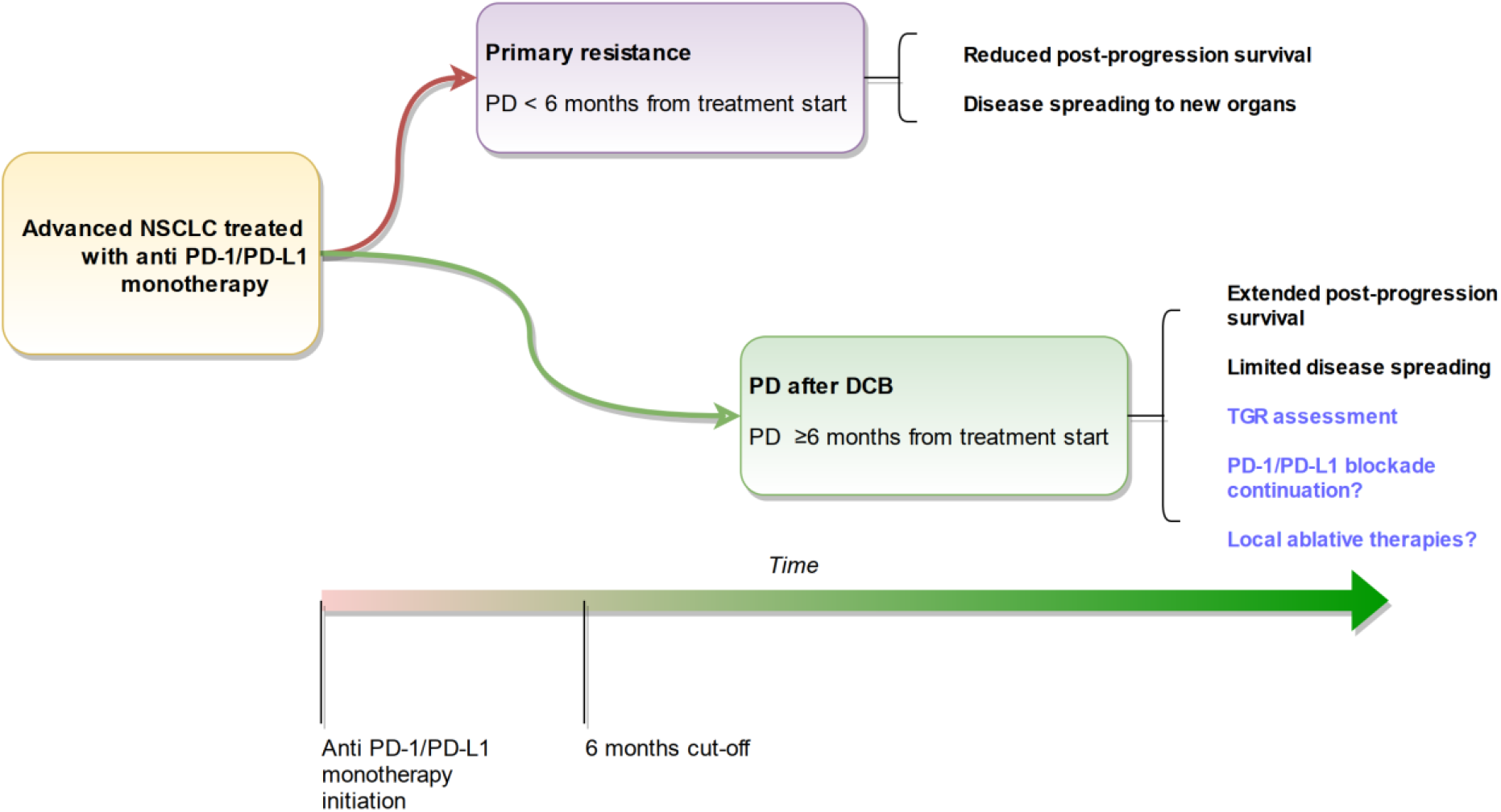
Summary of findings and perspectives.

## Data Availability

All data produced in the present study are available upon reasonable request to the authors

## Figures legends

**Supplementary Figure S1: Tumor Growth Rate model**

## Tables legends

**Supplementary Table S1: Patients with potential benefit of continuing PD-1/PD-L1 blockade beyond PD**. Highlighted lines are patients whose TGR assessed at PD was either null/negative, or inferior to TGR measured at baseline before ICI when both time points were available. Total = 19/43 patients.

**Supplementary Table S2: Oligoprogression characterization**. « Oligo PD lesions»: number of progressing lesions in OPD under PD-1/PD-L1 blockade; « New lesions PD2 »: presence of new lesions on « PD2 » imagery following first PD; « Interval PD-PD2 »: time (months) elapsed between both imageries; « TGR oligo »: TGR calculated specifically on progressing lesions in assessable OPD patients.

**Supplementary Table S3: Subsequent treatment after primary resistance or PD after DCB with anti-PD-1/PD-L1**. « Locoregional treatment » : radiation therapy, local ablative therapies or surgery

## References

1. The Global Cancer Observatory. Cancer fact sheet - All cancers. https://gco.iarc.fr/today/data/factsheets/cancers/39-All-cancers-fact-sheet.pdf

2. Sacher AG, L. LW, Lau A, Earle CC, Leighl NB. Real-world chemotherapy treatment patterns in metastatic non-small cell lung cancer: Are patients undertreated? Cancer. 2015;121(15):2562–2569. doi:10.1002/cncr.29386

3. Borghaei H, Paz-Ares L, Horn L, et al. Nivolumab versus Docetaxel in Advanced Nonsquamous Non–Small-Cell Lung Cancer. New England Journal of Medicine. 2015;373(17):1627–1639. doi:10.1056/NEJMoa1507643

4. Reck M, Rodríguez-Abreu D, Robinson AG, et al. Pembrolizumab versus Chemotherapy for PD-L1–Positive Non–Small-Cell Lung Cancer. https://doi.org/10.1056/NEJMoa1606774. doi:10.1056/NEJMoa1606774

5. Research C for DE and. Approved Drugs - Nivolumab (Opdivo). Accessed July 6, 2021. http://wayback.archive-it.org/7993/20170111231639/ http://www.fda.gov/Drugs/InformationOnDrugs/ApprovedDrugs/ucm436566.htm

6. Research C for DE and. Approved Drugs - Pembrolizumab injection. Accessed July 6, 2021. http://wayback.archive-it.org/7993/20170111231626/ http://www.fda.gov/Drugs/InformationOnDrugs/ApprovedDrugs/ucm465650.htm

7. Research C for DE and. Approved Drugs - Atezolizumab (TECENTRIQ). Accessed July 7, 2021. http://wayback.archive-it.org/7993/20170111231550/ http://www.fda.gov/Drugs/InformationOnDrugs/ApprovedDrugs/ucm525780.htm

8. Hirsch L, Zitvogel L, Eggermont A, Marabelle A. PD-Loma: a cancer entity with a shared sensitivity to the PD-1/PD-L1 pathway blockade. Br J Cancer. 2019;120(1):3–5. doi:10.1038/s41416-018-0294-4

9. Jin Y, An X, Mao B, et al. Different syngeneic tumors show distinctive intrinsic tumor-immunity and mechanisms of actions (MOA) of anti-PD-1 treatment. Sci Rep. 2022;12(1):3278. doi:10.1038/s41598-022-07153-z

10. Kluger HM, Tawbi HA, Ascierto ML, et al. Defining tumor resistance to PD-1 pathway blockade: recommendations from the first meeting of the SITC Immunotherapy Resistance Taskforce. J Immunother Cancer. 2020;8(1):e000398. doi:10.1136/jitc-2019-000398

11. Carbone DP, Reck M, Paz-Ares L, et al. First-Line Nivolumab in Stage IV or Recurrent Non-Small-Cell Lung Cancer. N Engl J Med. 2017;376(25):2415–2426. doi:10.1056/NEJMoa1613493

12. Brahmer J, Reckamp KL, Baas P, et al. Nivolumab versus Docetaxel in Advanced Squamous-Cell Non-Small-Cell Lung Cancer. N Engl J Med. 2015;373(2):123–135. doi:10.1056/NEJMoa1504627

13. Pathak R, Pharaon RR, Mohanty A, Villaflor VM, Salgia R, Massarelli E. Acquired Resistance to PD-1/PD-L1 Blockade in Lung Cancer: Mechanisms and Patterns of Failure. Cancers (Basel). 2020;12(12). doi:10.3390/cancers12123851

14. Spagnolo F, Boutros A, Cecchi F, Croce E, Tanda ET, Queirolo P. Treatment beyond progression with anti-PD-1/PD-L1 based regimens in advanced solid tumors: a systematic review. BMC Cancer. 2021;21(1):425. doi:10.1186/s12885-021-08165-0

15. Tian T, Yu M, Yu Y, et al. Immune checkpoint inhibitor (ICI)-based treatment beyond progression with prior immunotherapy in patients with stage IV non-small cell lung cancer: a retrospective study. Transl Lung Cancer Res. 2022;11(6):1027–1037. doi:10.21037/tlcr-22-376

16. Champiat S, Dercle L, Ammari S, et al. Hyperprogressive Disease Is a New Pattern of Progression in Cancer Patients Treated by Anti-PD-1/PD-L1. Clinical Cancer Research. 2017;23(8):1920–1928. doi:10.1158/1078-0432.CCR-16-1741

17. Berge DMHJ ten, Hurkmans DP, Besten I den, et al. Tumour growth rate as a tool for response evaluation during PD-1 treatment for non-small cell lung cancer: a retrospective analysis. ERJ Open Research. 2019;5(4). doi:10.1183/23120541.00179-2019

18. Tumati V, Iyengar P. The current state of oligometastatic and oligoprogressive non-small cell lung cancer. J Thorac Dis. 2018;10(Suppl 21):S2537–S2544. doi:10.21037/jtd.2018.07.19

19. Kim C, Hoang CD, Kesarwala AH, Schrump DS, Guha U, Rajan A. Role of Local Ablative Therapy in Patients with Oligometastatic and Oligoprogressive Non–Small Cell Lung Cancer. Journal of Thoracic Oncology. 2017;12(2):179–193. doi:10.1016/j.jtho.2016.10.012

20. Harris PA, Taylor R, Thielke R, Payne J, Gonzalez N, Conde JG. Research electronic data capture (REDCap)—A metadata-driven methodology and workflow process for providing translational research informatics support. Journal of Biomedical Informatics. 2009;42(2):377–381. doi:10.1016/j.jbi.2008.08.010

21. Harris PA, Taylor R, Minor BL, et al. The REDCap consortium: Building an international community of software platform partners. Journal of Biomedical Informatics. 2019;95:103208. doi:10.1016/j.jbi.2019.103208

22. Heo JY, Yoo SH, Suh KJ, et al. Clinical pattern of failure after a durable response to immune check inhibitors in non-small cell lung cancer patients. Sci Rep. 2021;11(1):2514. doi:10.1038/s41598-021-81666-x

23. Gettinger SN, Wurtz A, Goldberg SB, et al. Clinical Features and Management of Acquired Resistance to PD-1 Axis Inhibitors in 26 Patients With Advanced Non-Small Cell Lung Cancer. J Thorac Oncol. 2018;13(6):831–839. doi:10.1016/j.jtho.2018.03.008

24. Hosoya K, Fujimoto D, Morimoto T, et al. Clinical factors associated with shorter durable response, and patterns of acquired resistance to first-line pembrolizumab monotherapy in PD-L1-positive non-small-cell lung cancer patients: a retrospective multicenter study. BMC Cancer. 2021;21(1):346. doi:10.1186/s12885-021-08048-4

25. Schoenfeld AJ, Rizvi HA, Memon D, et al. Systemic and Oligo-Acquired Resistance to PD-(L)1 Blockade in Lung Cancer. Clinical Cancer Research. 2022;28(17):3797–3803. doi:10.1158/1078-0432.CCR-22-0657

26. Kagawa Y, Furuta H, Uemura T, et al. Efficacy of local therapy for oligoprogressive disease after programmed cell death 1 blockade in advanced non-small cell lung cancer. Cancer Sci. 2020;111(12):4442–4452. doi:10.1111/cas.14605

27. Laurie SA, Banerji S, Blais N, et al. Canadian consensus: oligoprogressive, pseudoprogressive, and oligometastatic non-small-cell lung cancer. Curr Oncol. 2019;26(1):e81–e93. doi:10.3747/co.26.4116

28. Prelaj A, Pircher CC, Massa G, et al. Beyond First-Line Immunotherapy: Potential Therapeutic Strategies Based on Different Pattern Progressions: Oligo and Systemic Progression. Cancers (Basel). 2021;13(6):1300. doi:10.3390/cancers13061300

29. Hu X, Li H, Kang X, et al. First-Line Tyrosine Kinase Inhibitors Combined With Local Consolidative Radiation Therapy for Elderly Patients With Oligometastatic Non-Small Cell Lung Cancer Harboring EGFR Activating Mutations. Frontiers in Oncology. 2022;12. Accessed May 28, 2022. https://www.frontiersin.org/article/10.3389/fonc.2022.766066

30. Lahmar J, Facchinetti F, Koscielny S, et al. Effect of tumor growth rate (TGR) on response patterns of checkpoint inhibitors in non-small cell lung cancer (NSCLC). JCO. 2016;34(15_suppl):9034–9034. doi:10.1200/JCO.2016.34.15_suppl.9034

31. He L na, Zhang X, Li H, et al. Pre-Treatment Tumor Growth Rate Predicts Clinical Outcomes of Patients With Advanced Non-Small Cell Lung Cancer Undergoing Anti-PD-1/PD-L1 Therapy. Front Oncol. 2021;10. doi:10.3389/fonc.2020.621329

32. Zou R, Wang Y, Ye F, Zhang X, Wang M, Cui S. Mechanisms of primary and acquired resistance to PD-1/PD-L1 blockade and the emerging role of gut microbiome. Clin Transl Oncol. Published online May 17, 2021. doi:10.1007/s12094-021-02637-2

33. Weiss SA, Sznol M. Resistance mechanisms to checkpoint inhibitors. Current Opinion in Immunology. 2021;69:47–55. doi:10.1016/j.coi.2021.02.001

34. Liu Y, Gu Y, Han Y, et al. Tumor Exosomal RNAs Promote Lung Pre-metastatic Niche Formation by Activating Alveolar Epithelial TLR3 to Recruit Neutrophils. Cancer Cell. 2016;30(2):243–256. doi:10.1016/j.ccell.2016.06.021

35. Engblom C, Pfirschke C, Zilionis R, et al. Osteoblasts remotely supply lung tumors with cancer-promoting SiglecFhigh neutrophils. Science. 2017;358(6367):eaal5081. doi:10.1126/science.aal5081

36. Altorki NK, Markowitz GJ, Gao D, et al. The lung microenvironment: an important regulator of tumour growth and metastasis. Nat Rev Cancer. 2019;19(1):9–31. doi:10.1038/s41568-018-0081-9

37. Shah S, Wood K, Labadie B, et al. Clinical and molecular features of innate and acquired resistance to anti-PD-1/PD-L1 therapy in lung cancer. Oncotarget. 2018;9(4):4375–4384. doi:10.18632/oncotarget.23315

38. Schoenfeld AJ, Antonia SJ, Awad MM, et al. Clinical definition of acquired resistance to immunotherapy in patients with metastatic non-small-cell lung cancer. Annals of Oncology. 2021;32(12):1597–1607. doi:10.1016/j.annonc.2021.08.2151

39. Ferté C, Fernandez M, Hollebecque A, et al. Tumor growth rate is an early indicator of antitumor drug activity in phase I clinical trials. Clin Cancer Res. 2014;20(1):246–252. doi:10.1158/1078-0432.CCR-13-2098

