## Supplementary Figure S1 for "Post-progression survival in advanced non-small cell lung cancer treated with anti-PD-1/PDL-1 monotherapy: progression after durable clinical benefit versus primary resistance"

**Cancer model:**

- *Virtual tumor sphere of volume « V » and radius « R »= D/2*
- *D = sum of target lesions per RECIST 1.1*
- *Tumor sphere growth: exponential function of time « t »*

$$V = 4\,\pi\,R^3/3$$

$$V_t = V_0 \exp(TG.t)$$

$$TG = 3\log(Dt/D0)/t$$

$$TGR = 100\,(\exp(TG) - 1)$$

*Growth rate, in %/month, of the volume of the global virtual tumor sphere*

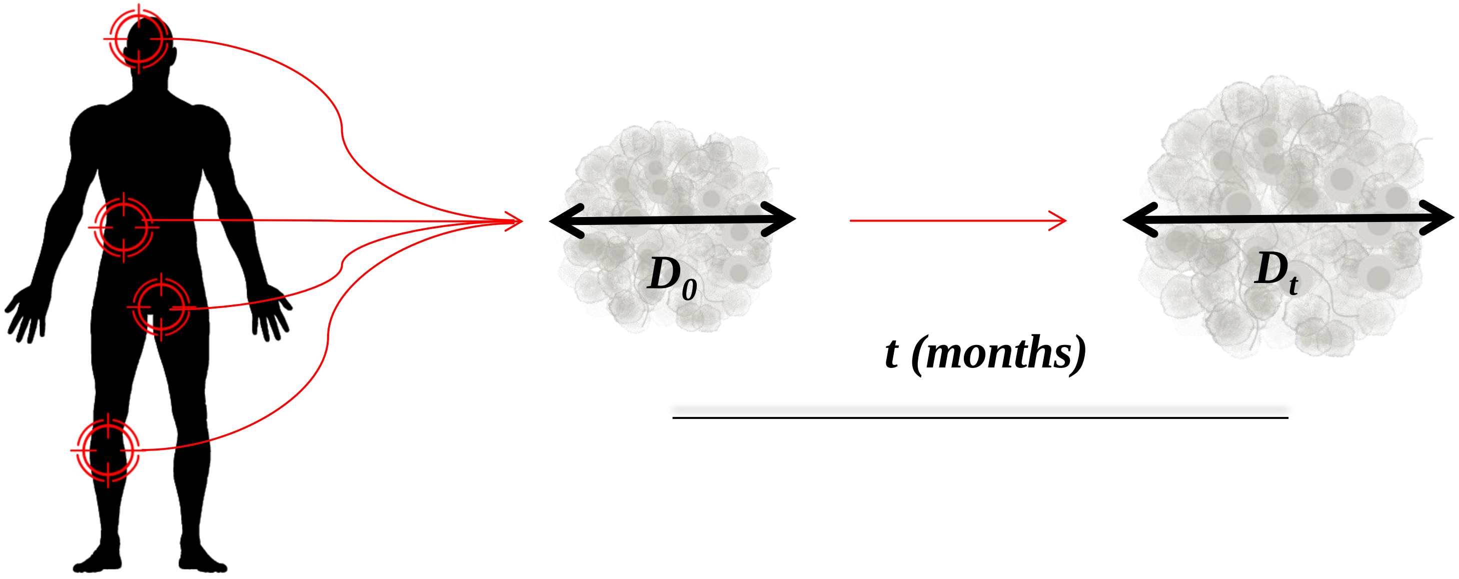
