## Supplementary Tables S1, S2 and S3 for "Post-progression survival in advanced non-small cell lung cancer treated with anti-PD-1/PDL-1 monotherapy: progression after durable clinical benefit versus primary resistance"

| **Record ID** | **TGR at baseline** | **TGR at PD** | **(TGR at PD) – (TGR at baseline)** |
| --- | --- | --- | --- |
| 20 | 0 | 0 | 0 |
| 34 | 0 | 0 | 0 |
| 118 | 12.388174267142 | 1.7414546411449683 | -10.646719625997 |
| 131 | 12.224674712953787 | 0 | -12.224674712953787 |
| 100 | 159.09177649222505 | 0 | -159.09177649222505 |
| 14 | 20.64531550717712 | 0 | -20.64531550717712 |
| 123 | 24.339482530031532 | 2.3903980877528763 | -21.949084442278654 |
| 82 | 45.495517688939536 | 22.62471283020291 | -22.870804858736626 |
| 99 | 263.9109874292884 | -13.598883758907165 | -277.5098711881956 |
| 143 | 50.141727065929366 | 17.897769952690457 | -32.243957113238906 |
| 139 | 77.40422359922444 | 44.235411086081825 | -33.16881251314261 |
| 96 | 38.83106847625 | 0 | -38.83106847625 |
| 85 | 53.82943084646092 | 0 | -53.82943084646092 |
| 125 | 543.5177915994036 | 0 | -543.5177915994036 |
| 113 | 3.9356266368534 | 4.3653138036101 | 0.42968716675673 |
| 51 | 10.67980381222795 | 20.852463043851486 | 10.172659231623536 |
| 32 | -9.149241654065154 | 4.358268100050711 | 13.507509754115866 |
| 65 | 0 | 20.13360462037759 | 20.13360462037759 |
| 138 | 11.730493284870281 | 15.951503891896234 | 4.221010607025953 |
| 130 | 0 | *NA* | *NA* |
| 61 | *NA* | 20.31635783321264 | *NA* |
| 107 | *NA* | 0 | *NA* |
| 135 | *NA* | *NA* | *NA* |
| 16 | 10.93009117412 | *NA* | *NA* |
| 95 | *NA* | *NA* | *NA* |
| 25 | *NA* | 27.26758888817349 | *NA* |
| 78 | *NA* | 13.979438856195 | *NA* |
| 38 | *NA* | -7.0184637492031 | *NA* |
| 59 | *NA* | 0 | *NA* |
| 137 | *NA* | *NA* | *NA* |
| 89 | *NA* | *NA* | *NA* |
| 92 | *NA* | 8.031584359335241 | *NA* |
| 115 | *NA* | 0 | *NA* |
| 116 | *NA* | -35.12330378991061 | *NA* |
| 56 | 36.593193844281 | *NA* | *NA* |
| 136 | *NA* | *NA* | *NA* |
| 142 | *NA* | 91.11066136359516 | *NA* |
| 128 | 4.174579567361 | *NA* | *NA* |
| 127 | 5.121696112977525 | *NA* | *NA* |
| 67 | *NA* | *NA* | *NA* |
| 122 | *NA* | 45.91109068426906 | *NA* |
| 129 | *NA* | *NA* | *NA* |
| 63 | *NA* | *NA* | *NA* |
|  |  |  | **(PD – baseline ≤ 0) OR (PD ≤ 0):**  n= 19 |

Supplementary Table S1: Patients with potential benefit of continuing PD-1/PD-L1 blockade beyond PD. Highlighted lines are patients whose TGR assessed at PD was either null/negative, or inferior to TGR measured at baseline before ICI when both time points were available. Total = 19/43 patients.

| **Oligo PD lesions** | **New lesions at PD2** | **Interval PD-PD2 (months)** | **TGR oligo (%/month)** |
| --- | --- | --- | --- |
| 1 | No | 2,73 | 0,00 |
| 1 | No | 2,79 | 2,93 |
| 2 | No | 3,35 | 4,80 |
| 2 | No | 2,30 | 7,33 |
| 1 | No | 1,28 | 11,53 |
| 1 | No | 3,48 | 12,12 |
| 2 | No | 2,60 | 18,93 |
| 1 | No | 3,88 | 20,07 |
| 2 | No | 1,28 | 20,32 |
| 2 | No | 2,30 | 20,85 |
| 1 | No | 1,54 | 22,24 |
| 1 | No | 3,88 | 23,00 |
| 1 | No | 1,12 | 26,12 |
| 1 | No | 2,00 | 31,16 |
| 1 | No | 1,15 | 32,12 |
| 1 | No | 2,07 | 32,71 |
| 2 | No | 2,33 | 34,20 |
| 2 | No | 2,76 | 35,04 |
| 1 | No | 2,07 | 35,15 |
| 1 | No | 1,91 | 41,68 |
| 1 | No | 3,61 | 54,62 |
| 1 | No | 2,33 | 58,30 |
| 2 | No | 1,77 | 145,31 |
| 2 | No | 2,23 | 160,37 |
| 1 | Yes | 2,33 | *NA* |
| 2 | Yes | 2,99 | *NA* |
| 1 | Yes | 2,46 | *NA* |
| 1 | Yes | 2,04 | *NA* |
| 3 | *NA* | *NA* | *NA* |
| 3 | *NA* | *NA* | *NA* |
| 1 | *NA* | *NA* | *NA* |
| 2 | *NA* | *NA* | *NA* |
|  |  | Mean PD-PD2 | Median TGR oligo |
|  |  | 2,378165869 | 24,56 (IQR 17,84) |

Supplementary Table S2: Oligoprogression characterization. « Oligo PD lesions» : number of progressing lesions in OPD under PD-1/PD-L1 blockade ; « New lesions PD2 » : presence of new lesions on « PD2 » imagery following first PD ; « Interval PD-PD2 » : time (months) elapsed between both imageries ; « TGR oligo » : TGR calculated specifically on progressing lesions in assessable OPD patient.

|  |  | Primary resistance (N=105) | PD after DCB  (N=43) |
| --- | --- | --- | --- |
| No treatment |  | 38 | 6 |
| Subsequent treatment | Anti-PD-1/PD-L1monotherapy continuation + locoregional treatment | 2 | 3 |
|  | Anti-PD-1/PD-L1 monotherapy rechallenge after toxicity | 0 | 3 |
|  | Anti-PD-1/PD-L1monotherapy continuation despite PD | 9 | 5 |
|  | Locoregional treatment alone | 1 | 2 |
|  | New systemic therapy | 55 | 24 |
|  | Platine doublet +or- bevacizumab | 11 | 3 |
|  | Taxane +or- bevacizumab | 25 | 6 |
|  | Tyrosine kinase inhibitor | 8 | 3 |
|  | Pemetrexed | 5 | 7 |
|  | Vinorelbine | 3 | 0 |
|  | Gemcitabine | 3 | 5 |

Supplementary Table S3: Subsequent treatment after primary resistance or PD after DCB with anti-PD-1/PD-L1. « Locoregional treatment » : radiation therapy, local ablative therapies or surgery
